# The cumulative impact of clinical risk on brain networks and associations with executive function impairments in adolescents with congenital heart disease

**DOI:** 10.1101/2023.12.06.23299632

**Authors:** Melanie Ehrler, Anna Speckert, Oliver Kretschmar, Ruth Tuura O’Gorman, Beatrice Latal, Andras Jakab

**Affiliations:** Child Development Center, University Children’s Hospital Zurich, Switzerland; Children’s Research Centre, University Children’s Hospital Zurich, Switzerland; University Research Priority Program (URPP), Adaptive Brain Circuits in Development and Learning (AdaBD), University of Zurich, Switzerland; Center for MR Research, University Children’s Hospital Zurich, Switzerland; Pediatric Cardiology, Pediatric Heart Center, Department of Surgery, University Children’s Hospital Zurich, Switzerland

## Abstract

Congenital heart disease (CHD) is known to negatively affect brain development. Individual clinical factors have been identified to contribute to this detrimental effect, however, the cumulative effect of these factors on brain development and neurodevelopmental outcomes are not fully understood. Our study utilized structural brain connectomics as an indicator of brain development to investigate the potential combined impact of clinical risk factors and family-environmental factors in adolescents with CHD.

We developed a cumulative clinical risk (CCR) score by summing up binary risk factors (neonatal, cardiac, neurologic) based on clinically relevant thresholds and further investigated the role of family-environmental factors (parental education, parental mental health, and family function). Brain development in 53 adolescents with CHD who underwent infant open-heart surgery, and 75 healthy controls was studied by diffusion MRI connectomic analysis and neuropsychological assessments. The CCR score explains a great variability of the structural brain connectome. A higher CCR score was associated with lower network segregation, edge strength, and executive functioning. Edge strength was particularly reduced in an inter-frontal and fronto-parietal-thalamic network. There was no association with family-environmental factors. Poorer executive functioning was associated with lower network integration and segregation.

Our results provide evidence for persisting alterations of network connectivity in adolescents with CHD – particularly in those patients who face a cumulative exposure to multiple clinical risk over time. Quantifying the cumulative load of risk early in life, may help to better predict trajectories of brain development in order to identify and support the most vulnerable patients as early as possible.

## 1. Introduction

Congenital heart disease (CHD) is the most common congenital malformation, affecting approximately 1% of all newborns.^1^ About one third of these children require infant cardiopulmonary bypass (CPB) surgery. Improved surgical and medical management over the past decades has led to reduced mortality and morbidity and consequently to a shift of attention towards long-term outcome of patients with CHD. Despite favorable cardiac outcome in the majority of these patients, neurodevelopmental impairments are frequently observed across the lifespan and have been recognized as the most prevalent non-cardiac comorbidity in CHD.^2^ A meta-analysis demonstrated an increased risk for mild intellectual and executive function impairments in children and adolescents with CHD who underwent CPB surgery. Importantly, all executive function domains were similarly affected without a specific pattern. These domains included working memory, inhibition, cognitive flexibility, fluency and planning.^3^ Importantly, there is evidence that executive function problems persist into adulthood.^4^ These difficulties have been associated with lower quality of life^5^, higher need for educational support^6^, and co-existing socio-behavioral difficulties and anxiety symptoms^7^. Understanding the neuronal alterations that lead to neurodevelopmental impairments is essential, given their impact on everyday functioning.

Neuroimaging studies have elucidated brain alterations in patients with CHD compared to healthy individuals. Fetal studies showed that brain development is already altered intrauterine and brain developmental trajectories continue to be impaired. Fetuses with complex CHD exhibit reduced brain growth and delayed cortical development, at least partly comparable to preterm babies. ^8–10^ Brain growth remains altered from the pre-to postnatal period^11,12^, and is associated with an increased risk for postnatal white matter injury.^9^ Studies using diffusion weighted MRI (dMRI) in neonates with CHD have demonstrated altered integrity of white matter tracts^13–18^ and structural network connectivity^19–21^. According to these studies, brain development in CHD is characterized by altered network-level properties of the structural connectome. These connectomic findings indicate a less efficient neuronal communication and delayed maturation of structural networks.^19–21^ A recent study found that lower preoperative network integration was associated with a severity score based on alterations of fetal substrate delivery (i.e., how much the cardiac malformation alters typical fetal circulation). In contrast, postoperative network integration was associated with perioperative factors (i.e., longer bypass and ECMO time).^22^ To underline the clinical relevance of these alterations, One study demonstrated that parent-reported externalizing problems at the age of two years were associated with lower edge strength of a frontal-limbic network assessed preoperatively.^23^

Currently, there is limited evidence on whether the altered network connectivity observed in neonates persists throughout the lifespan. While a body of research has shown persisting alterations of microstructural integrity in several white matter tracts of adolescents and adults with CHD^24–28^, only two studies have investigated structural network connectivity beyond the neonatal period. One study in adolescents with transposition of the great arteries (TGA) showed increased network segregation and associations with lower cognitive performance in patients relative to healthy controls.^29^ A study in 13 children with single-ventricle physiology and 13 controls did not detect any group differences in network integration or network segregation.^30^

Across the reported dMRI studies, between-study variability is considerably high likely due to the heterogeneity of included patients (e.g., different CHD diagnoses with varying clinical risk profile) and imaging approaches (e.g., different dMRI sequences and processing pipelines). Consequently, dMRI alone can currently not be used as a biomarker for aberrant brain developmental trajectories in individuals with CHD. More emphasis needs to be placed on characterizing specifically vulnerable patients using marker of CHD severity and other clinical risk factors. Moreover, while family-environmental factors have been identified as strong determinants of neurodevelopment and behavior in this group ^31,32^, their influence on brain development remains unexplored. Studies in typically-developing individuals found that environmental factors, such as socioeconomic status impact brain development. It has been suggested that high socioeconomic background is related to extended cortical development and increased segregation of functional networks in adolescence which indicates refinement and efficiency of neuronal processes.^33^ The role of socioeconomic background and other family-related factors for brain development in patients with CHD is sought to be investigated. In addition, studies investigating associations between alteration of structural brain networks and neurodevelopmental impairments are needed to deepen our understanding of the neuronal processes contributing to the observed developmental problems in these patients.

Therefore, we aimed to identify alterations of network connectivity measures derived from dMRI in adolescents with CHD relative to healthy controls. Network connectivity measures of interest included a) network integration (measured by global efficiency), b) network segregation (measured by local efficiency) and network edge strength. Further, we aimed to investigate if these network connectivity measures were associated with the patients’ cumulative load of clinical risk factors and with family-environmental factors. Lastly, we aimed to determine the relationship between network connectivity measures and executive function impairments.

## 2. Materials and Methods

### 2.1. Sample and study design

The data used for this analysis is derived from a prospective cohort study (Teen Heart Study ^59^). This study investigated neurodevelopmental outcomes and cerebral MRI in adolescents with CHD and was conducted at the University Children’s Hospital of Zurich between 2019 and 2021.

The study included patients with CHD who underwent CPB surgery between 2004 and 2012 at the University Children’s Hospital Zurich. Patients were eligible if they underwent CPB surgery within their first year of life, have not been diagnosed with a genetic disease or a syndromal disorder and were between 10 and 15 years of age at the time of assessment. Of 178 eligible patients, 100 patients participated in the current study (participation rate: 56%). A control group of 104 healthy adolescents was cross-sectionally recruited. Inclusion criteria were birth after 36;0 weeks of gestation and no diagnosis of a neurological or developmental disorder (i.e., learning disorder or attention deficit hyperactivity disorder). The recruitment procedure is displayed in Supplementary Figure 1. The study was approved by the ethics committee of the Canton of Zurich, Switzerland (KEK 2019–00035). Written informed consent was obtained prior to participation from the participant’s legal guardian and from participants who were 14 years or older. De-identified data and statistical scripts supporting our findings can be shared specifically with other groups upon reasonable request. Patient data cannot be made openly available due to patient privacy reasons.

### 2.2. Clinical risk factors and family-environmental factors

Clinical risk factors were prospectively collected from patients’ clinical records. A cumulative clinical risk score was created from the sum of dichotomized risk factors (1=risk, 0=no risk). The selection of risk factors was based on a literature search as previously reported^31^ and the definition of risk thresholds (i.e., dichotomizing) were done in a consensus discussion based on clinical considerations between OK (cardiologist) BL (developmental pediatrician) and ME (psychologist). The following dichotomized risk factors were selected: 1) Single-ventricle physiology, 2) cyanotic physiology, 3) preterm birth (<37 weeks of gestation), 4) low birthweight (<5^th^ percentile, adjusted for gestational age and sex), 5) low head circumference at birth (<5^th^ percentile, adjusted for gestational age and sex), 6) preoperative O_2_ saturation <70%, 7) >1 open-heart surgery, 8) deep hypothermia during first open-heart surgery (<28°C), 9) extracorporeal circulation time >120min, 10) >2 weeks of intensive care unit stay following the first surgery, 11) having been on ECMO, 12) having had a stroke identified on routine and/or research cerebral MRI, 13) having had a clinically noticeable seizure, 14) need for cardiac medication at time of assessment (as a measure for impaired heart function). The *cumulative clinical risk score* (CCR score) could range from 0 to 14.

The following family-environmental factors were assessed at the time of assessment: parental education, parental mental health and family functioning. Parental education was calculated by the sum of maternal and paternal education, each assessed on a 6-point scale (1 = no high-school degree, 2 = high-school degree, 3 = apprenticeship, 4 = higher diploma for craftsmen or craftswomen, 5 = advanced diploma of higher education, 6 = university degree).^60^ Maternal mental health was measured with the Brief Symptom Inventory (BSI-18), a standardized questionnaire that was filled in by the mothers and assesses symptoms of depression, anxiety and somatization. An age and sex-adjusted total score was calculated based on representative German norms.^61^ Family functioning was assessed with the Family Relationship Index (FRI), a standardized questionnaire that was filled in by the mothers and assesses the domains cohesion (degree of commitment, help and support), expressiveness (degree of expressed feelings and open interaction) and conflict (degree of expressed anger, and aggression). A total score for overall quality of family functioning was calculated by the sum of the three subdomains, after reversal of the subdomain conflict. Higher scores reflect better quality of family functioning.^62^

### 2.3. Executive function performance

Executive functions were investigated with an extensive standardized neuropsychological test battery assessing working memory, inhibition, cognitive flexibility, verbal and design fluency, and problem solving.^59^ Details about the neuropsychological test measures of executive functions are displayed in Table 1. A standardized summary score for executive function performance was built based on these tests. Please refer to the Supplementary Text 1, Supplementary Table 1 and 2 for further information.

**Table 1:** Neuropsychological test battery to assess executive functions performance.

| **Executive function domains** | **Neuropsychological test** | **Test measurement** |
| --- | --- | --- |
| Working memory | Digit Span forward & backward (WISC-IV) | Number of correct items |
|  | Letter-number sequencing (WISC-IV) | Number of correct items |
|  | Corsi Block Tapping Test (Corsi) | Number of correct items |
| Inhibition | Subtest Interference, Colour Word Interference Task (D-KEFS) | Completion time |
|  | Go/NoGo (TAP) | Number of commission errors |
| Cognitive flexibility | Subtest Letter-Number-Switching, Trail Making Task (D-KEFS) | Completion time |
|  | TAP Flexibility (TAP) | Median reaction time |
| Fluency | Subtests s-words and animals (RWT) | Number of correct items |
|  | Subtest filled-dots-only, Design Fluency Test (D-KEFS) | Number of correct items |
| Planning | Tower Task (D-KEFS) | Total achievement score |

*Note.* WISC-IV = German version of the Wechsler Intelligence Scale for Children Fourth Edition <sup>36</sup>, Corsi <sup>37</sup>, D-KEFS = Delis-Kaplan
Executive Function System <sup>38</sup>, TAP = Test of Attentional Performance <sup>39</sup>, RWT = Regensburger verbal fluency Test <sup>40</sup>

### 2.4. Magnetic resonance imaging (MRI) acquisition

Cerebral MRI was performed on a 3 Tesla whole-body system (GE 3T MR750 MRI scanner, GE Healthcare, Milwaukee, WI). A diffusion imaging sequence and a three-dimensional spoiled gradient echo (SPGR) pulse sequence were acquired to estimate structural network connectivity.

MR acquisition parameters, diffusion weighted imaging processing and structural network construction is detailed in Supplementary Text 2 to 4. In summary, weighted, undirected structural connectivity networks were formed based on whole-brain probabilistic tractography resulting in 162×162 matrices where edges correspond to the sum of weighted streamlines (SIFT2*µ) and nodes correspond to the AAL3v1 labels.

### 2.5. Network connectivity measures

#### 2.5.1. Graph theory measures

The following graph theory measures were estimated with the Brain Connectivity Toolbox (BCT^68^) in MATLAB version R2022b to quantify the topological organization of the brain networks. Threshold-free *network density* was measured as the observed edges relative to all possible edges.^69^ The ‘density_und’ function in BCT was used. Network integration was quantified by *global efficiency*, which was defined as the inverse of the average shortest path length between nodes.^70^ Network segregation was quantified by mean *local efficiency*, which was defined as the inverse of the average shortest path length connecting all neighbors of an individual node, averaged across all nodes. Cost thresholding was applied to account for variance in network density between subjects. As the choice of cost threshold may be arbitrary, an iterative approach was used with a set of thresholds from 0.05 to 0.46 increasing in steps of 0.01. The upper limit (τ_max_=0.46) corresponds to the lowest possible network density across the study population. For cost thresholding, the function ‘threshold_proportional’ from BCT was used. Global efficiency and local efficiency were calculated at each cost thresholding level with the function ‘efficiency_wei’ from BCT ^68^. Nodes were portioned into *core* regions (well-connected regions, strongly interconnected hubs) or *periphery* regions (weakly connected regions, brain periphery).^71^ A common core-periphery pattern was defined across the study sample by classifying those nodes which were identified as core or periphery, respectively, in ≥90% of all subjects. Other nodes were classified as *neither*. The coreness statistic was estimated by the degree of separation between core and periphery nodes as a measure of goodness of fit. The ‘core_periphery_dir’ function from BCT was used.

#### 2.5.2. Network based statistics (NBS)

The MATLAB toolbox Network based statistics (NBS) was used to test the associations between edge-wise structural connectivity and factors of interest (i.e., group differences, association with CCR score and executive function summary score, please refer to section 2.8 for details). Networks were thresholded at a cost level of τ_max_=0.46. A general linear model was used with 5000 permutations to determine significance. First, mass univariate testing was conducted for each edge with a *T* threshold of 3.0. Post hoc, analyses were repeated with a *T* threshold of 3.5 to identify the strongest components of the network. Second, all edges exceeding the *T* threshold were admitted to a set of supra-threshold connections. Then, topological clusters among the set of supra-threshold connections were identified. Connections for which the null hypothesis had been rejected were arranged in an interconnected configuration rather than being single/isolated connections. Third, family-wise error rate correction was conducted for each component using permutation testing. The component size corresponds to the total number of connections it comprises (*extent*). P values < 0.05 were considered significant.

### 2.6. Statistical analyses

Statistical analyses were performed in R version 4.2.2 ^72^ if not stated elsewise. Sample characteristics were compared between patients and controls with t-tests for normally distributed continuous data, Mann Whitney U-tests for non-normally distributed continuous or ordinal data, and Chi-squared test for categorical data. Missing values of clinical risk factors were imputed by chained equation to maintain statistical power to calculate the CCR score. The R package ‘mice’ was used with 1 imputed data set and 5 iterations^73^.

Network connectivity measures were first compared between patients and controls. To test for group effects on threshold-free network density, a linear regression model was estimated with network density as dependent variable, group as independent variable, and age and sex as covariates. To investigate global and local efficiency, a mass univariate approach was used. First, global efficiency was log transformed to achieve normal distributions of residuals for all subsequent analyses. Linear regression models were estimated separately for each threshold level (from 0.05 to 0.46) with threshold specific global efficiency as dependent variable, group as independent variable, and age and sex as covariates. Group effects were FDR corrected. The same procedure was repeated for local efficiency.

*Subnetwork edge strength* was identified by NBS. Edge-wise general linear models were estimated with edge strength as dependent variable, group as independent variable, and age and sex as covariates in NBS (see further details in section 2.7.2). Mean network strength of an identified subnetwork was calculated for each individual and compared between groups with a two-sampled, two-sided t-test. Identified networks were visualized with BrainNet Viewer in MATLAB and with circular graphs in R with the libraries ‘ggplot2’, ‘igraph’ and ‘ggraph’.

Linear regression models were estimated with the following network connectivity measures were considered as dependent variables in separate models: a) global efficiency (averaged across all cost thresholds), b) local efficiency (averaged across all cost thresholds) and c) mean subnetwork edge strength identified by NBS-derived group comparison. Independent variables were: 1) CCR score, 2) individual clinical risk factors, estimated in separate models due to potential collinearity (FDR corrected p-values are reported), 3) family-environmental factors, 4) executive function summery score. All models were corrected for age and sex.

Post hoc, subnetwork edge strength specifically linked to the CCR score was identified by NBS within the patient group. To do so, edge-wise general linear models were estimated with edge strength as dependent variable, the cumulative clinical risk score as independent variable, and age and sex as covariates with NBS (see further details in section 2.7.2). The same procedure was repeated for the executive function summary score as independent variable.

Post hoc, it was tested if the association between the CCR score and the executive function summary score was mediated by local efficiency using the R package ‘lavaan’.

All linear regression models were checked for normal distribution of residuals and collinearity of factors included.

## 3. Results

### 3.1. Study sample and characteristics

Of 204 subjects who participated in the study (100 patients, 104 controls), 156 underwent cerebral MRI (60 patients, 95 controls). Primary reasons for not undergoing cerebral MRI were implants (e.g., pacemaker, stents, clips), which were not approved for research MRI, or anxiety of the participant. In two individuals, the dMRI sequence was not performed. Participants were excluded due to artifacts from dental braces (n=20), movement artifacts in the dMRI sequence (n=2) and in the SPGR sequence (N=1), artifacts after eddy current correction (n=1), insufficient data registration (n=1), and a large cyst (n=1). Eventually, 128 subjects were included in the current analysis. Included subjects did not differ from excluded subjects in respect to age (*t*(*df*)=1.95(202),*p*=0.052), sex (*Χ*^2^(*df*)=0.02(1),*p*=0.889), parental education (*W*=3837,*p=*0.104), proportion of patients with a cyanotic CHD (*Χ*^2^(*df*)=2.07(1),*p*=0.150), and with an univentricular CHD (*Χ*^2^(*df*)=1.08(1),*p*=0.298).

A total of 53 patients (10% univentricular CHD, 66% cyanotic CHD) and 75 healthy controls were analyzed. Subjects were between 10.1 and 15.9 years of age. Detailed participant characteristics stratified by group are reported in Table 2. Patients were significantly older and had significantly lower parental education than healthy controls. There was no significant difference between patients and controls with respect to sex, gestational age, birth weight and head circumference at birth. Patients had significantly lower total IQ and lower executive function summary score relative to healthy controls (IQ [100±15]: patients’ mean = 100.7±12.2, controls’ mean = 111.1±9.6, *p*<0.001; executive function summary score [0±1]: patients’ mean=-1.0±1.1; controls’ mean=-0.1±1.0,*p*<0.001). Registration quality of dMRI data did not differ significantly between patients and controls (*W*=1987,*p*=0.998).

**Table 2:** Participant characteristics

| Sample characteristic | Patients<br>(n = 53) | Controls<br>(n = 75) | p-value |
| --- | --- | --- | --- |
| **Innate** |  |  |  |
| Age in years (M[sd]) | 13.6(1.1) | 12.9(1.4) | **0.003<sup>a</sup>** |
| Male sex (N[%]) | 34(64%) | 36(48%) | 0.104<sup>b</sup> |
| Parental education (Mdn[IQR]) | 8(6-9) | 10(8-12) | **<0.001<sup>c</sup>** |
| **Neonatal** |  |  |  |
| Gestational age at birth (Mdn[IQR]) | 39.6(38.4-40.4) | 39.6(38.2-40.2) | 0.568<sup>c</sup> |
| Birth weight, z-score (M[sd]) | -0.3(1.1) | -0.3(1.0) | 0.815<sup>a</sup> |
| Head circumference, z-score (M[sd]) | -0.5(1.1) | -0.3(1.2) | 0.514<sup>a</sup> |
| Univentricular CHD (N[%]) | 10(19%) | - |  |
| Cynaotic CHD (N[%]) | 35(66%) | - |  |
| **Perioperative time** |  |  |  |
| Age at first surgery, months (Mdn[IQR]) | 0.95(0.26-4.36) | - |  |
| Mean preoperative O2 saturation, % (Mdn[IQR]); < 70% (N[%]) | 89(80-95); 3 (6%) | - |  |
| ECC time during first CPB surgery in minutes (M[sd]); >120 minutes (N[%]) | 168(69); 42 (79%) | - |  |
| Lowest temperature during first CPB surgery in °C (Mdn[IQR]); <28°C (N[%]) | 30(26-32); 26 (49%) | - |  |
| Time on ICU after first CPB surgery in days (Mdn[IQR]); >14 days (N[%]) | 7(5-10); 7 (13%) | - |  |
| Total number of CPB surgeries (Mdn[IQR]); >1 CPB surgery (N[%]) | 1(1-2); 15 (28%) | - |  |
| **Further clinical course** |  |  |  |
| Critical neurological or cardiac event ever (N[%]) |  |  |  |
| ECMO | 1(2%) | - |  |
| Clinically relevant seizure (ever) | 5(9%) | - |  |
| Stroke identified on cerebral MRI (ever) | 2(4%) | - |  |
| Cardiac medication at time of study assessment (N[%]) | 8(15%) | - |  |

Note: CHD = congenital heart disease, ECC = extracorporeal circulation, ECMO = extracorporeal membrane oxygenation ICU = intensive care unit, CPB = cardiopulmonary bypass. SD = standard deviation, IQR = interquartile range. <sup>a</sup> two-sampled two-sided t-test, <sup>b</sup> two-sampled Chi-squared test, <sup>c</sup> two-sampled, two sided Mann-Whitney U test. Bold p-value reference to a significant group difference.

The T1 and T2 weighted images were reviewed for incidental findings by a radiologist and only minor findings were detected in 6 patients, no incidental findings were detected in controls (see Supplementary Text 5). As these findings did not significantly interfere with the white matter structure and to maintain a representative sample, these subjects were not excluded for further analyses.

For patients, the clinical risk factors are reported in Table 2 and dichotomized risk factors are reported in Table 3. The CCR score ranged from 0 to 11 (*Mdn*=3,*IQR*=2 to 4). The CCR score correlates significantly with the executive function summary score (*r*=-0.33,*p*=0.016).

**Table 3:** Dichotomized clinical risk factors of the patient group

| Dichotomized risk factors | N (% of 53 patients) |
| --- | --- |
| Univentricular CHD | 10 (19%) |
| Cyanotic CHD | 35 (66%) |
| Preterm birth (<37 weeks of gestation) | 3 (6%) |
| Birth weight <5th percentile | 7 (13%) |
| Head circumference at birth <5th percentile | 8 (15%) |
| Mean preoperative O2 saturation < 70% | 3 (6%) |
| ECC time during first CPB surgery >120 minutes | 42 (79%) |
| Lowest temperature during first CPB surgery <28°C | 26 (49%) |
| Time on ICU after first CPB surgery >14 days | 7 (13%) |
| >1 CPB surgery | 15 (28%) |
| ECMO (ever) | 1 (2%) |
| Clinically relevant seizure (ever) | 5 (9%) |
| Stroke identified on cerebral MRI (ever) | 2 (4%) |
| Needing cardiac medication (at time of assessment) | 8 (15%) |

*Note: CHD = congenital heart disease, ECC = extracorporeal circulation, ECMO = extracorporeal membrane oxygenation ICU = intensive care unit, CPB = cardiopulmonary bypass. To estimate the cumulative clinical risk score, missing values were estimated with imputation by chained equation. Missing values: head circumference at birth n = 4, mean preop saturation n = 2; cardiac medication n = 1.*

### 3.2. Differences of network connectivity measures between patients and healthy controls

Patients had significantly lower threshold-free network density compared to healthy controls (*β*(*CI-95*)=0.28(0.1–0.45),*p*=0.002). There were no significant differences between patients and controls in global efficiency at any cost threshold levels (Figure 1). We found consistently lower local efficiency in patients relative to controls from a cost threshold of 0.12 or higher after FDR correction (Figure 2). Also, local efficiency averaged across all cost threshold was significantly lower in patients compared to controls (*β*(*CI-95*)=0.2(0.23–0.59),*p=*0.027).

**Figure 1:**
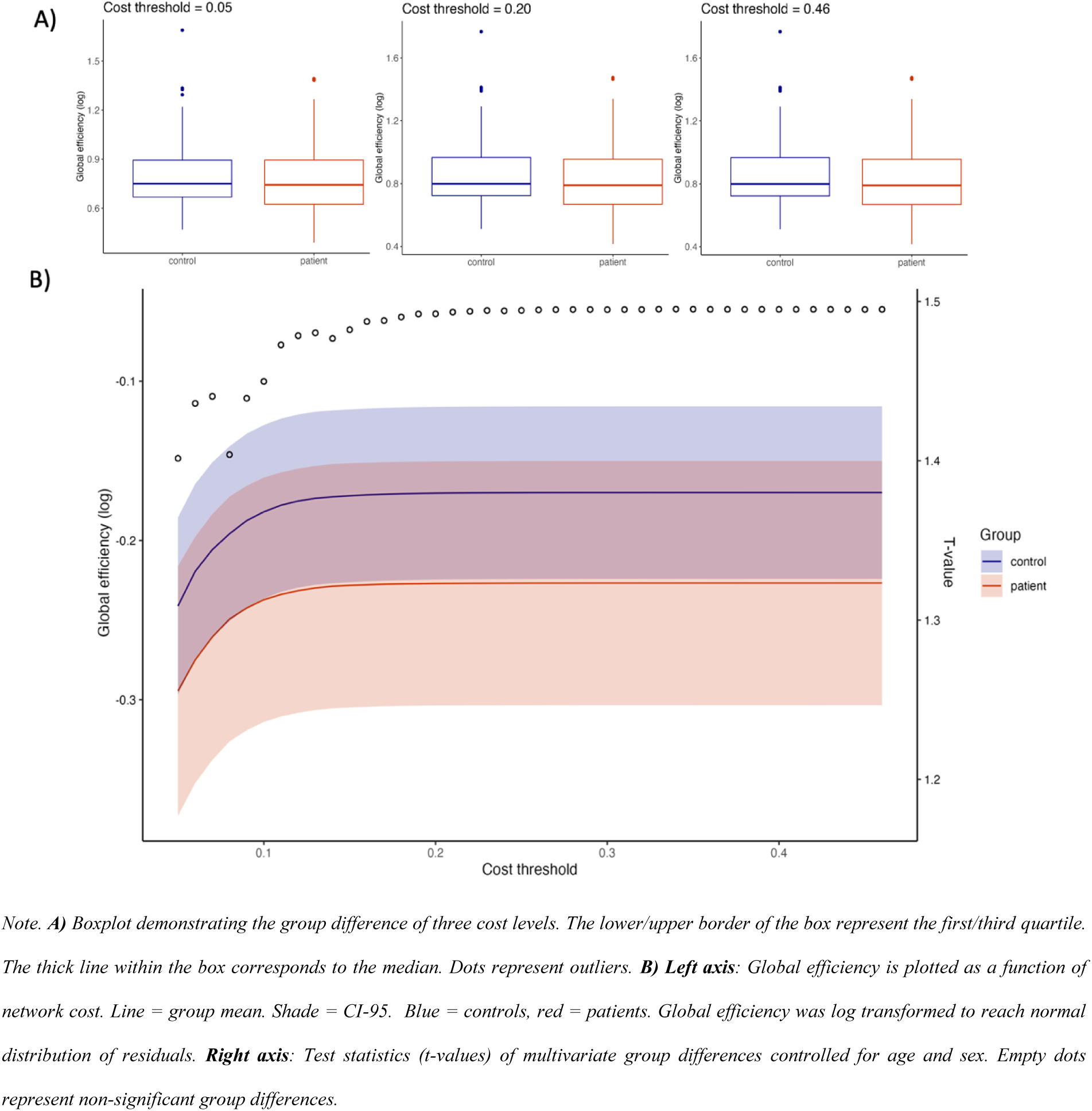
Differences between patients and controls in global efficiency at different cost threshold levels

**Figure 2:**
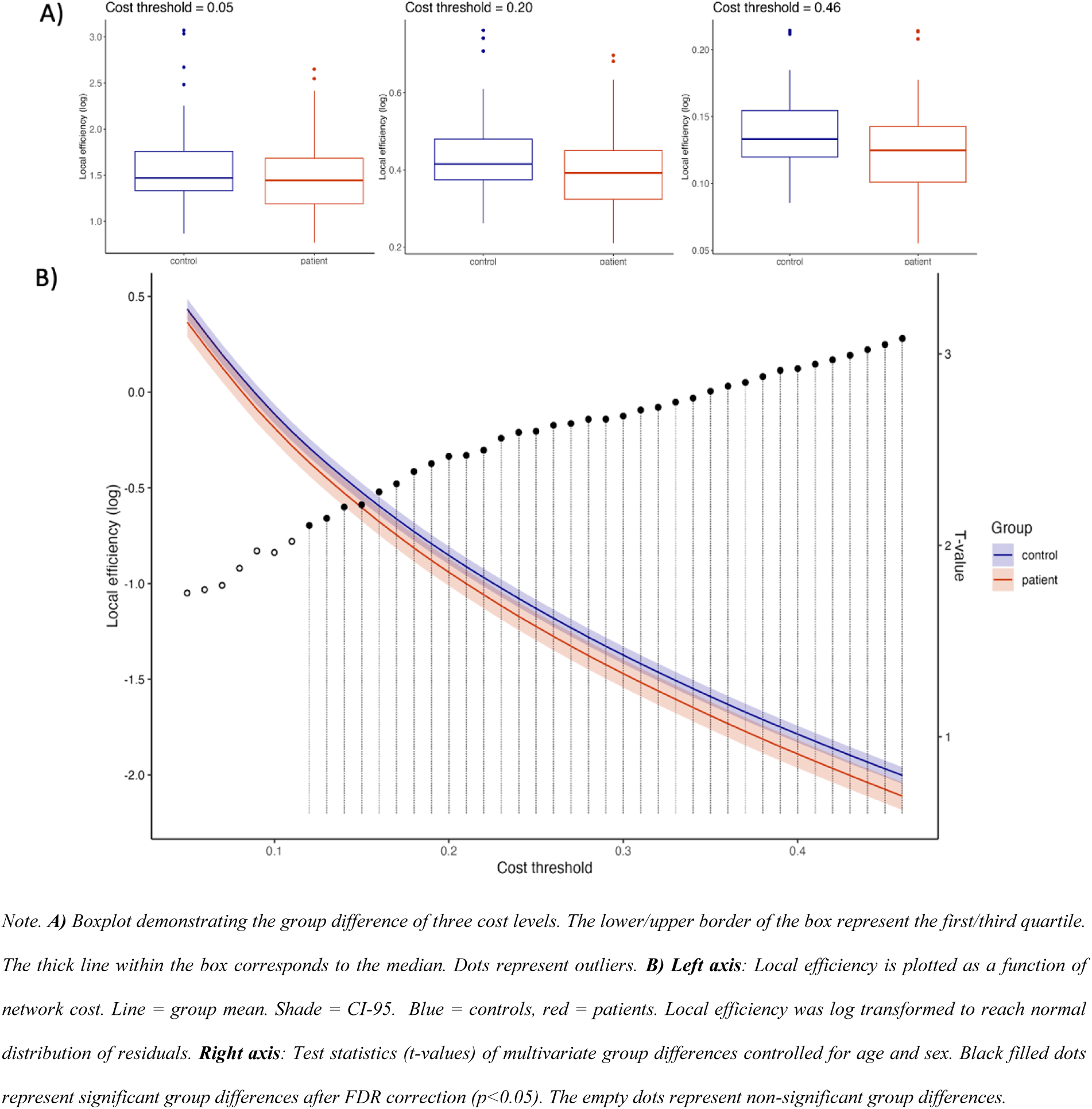
Differences between patients and controls in local efficiency at different cost threshold levels

Using NBS, we identified a dense subnetwork with lower network strength in patients compared controls. This network included 421 edges connecting 130 nodes (*p*<0.001, multiple comparison corrected with a T threshold of 3.0; see Figure 3 and Supplementary table 3 and 4). This analysis was repeated with a T threshold of 3.5 to identify the strongest component of the network in order to better describe the composition of nodes and edges involved (167 edges, 89 nodes, *p*<0.001; see supplementary figure 2 and table 3 and 5;). Qualitatively, the network involved all brain regions (frontal, parietal, temporal, and occipital lobes, limbic system, insula, deep grey matter, cerebellum) and comprised interconnections of the frontal lobe, interhemispheric connections and cortico-subcortical connections. The mean network strength of this subnetwork is called *subnetwork edge strength* in the following sections. The core-periphery analysis revealed that the majority of involved nodes were core regions. The thalamic nuclei were first classified as peripheral regions. However, when merging the atlas labels from all thalamic nuclei to one single region (as it has been done in NiBhroin et al., 2020^21^), the thalamus was classified as a core region as well. The coreness statistics was similar between patients and controls (*t*(*df*)=1.67(126),*p*=0.097). There was no subnetwork identified with higher network strength in patients relative to controls (*p=*0.127).

**Figure 3:**
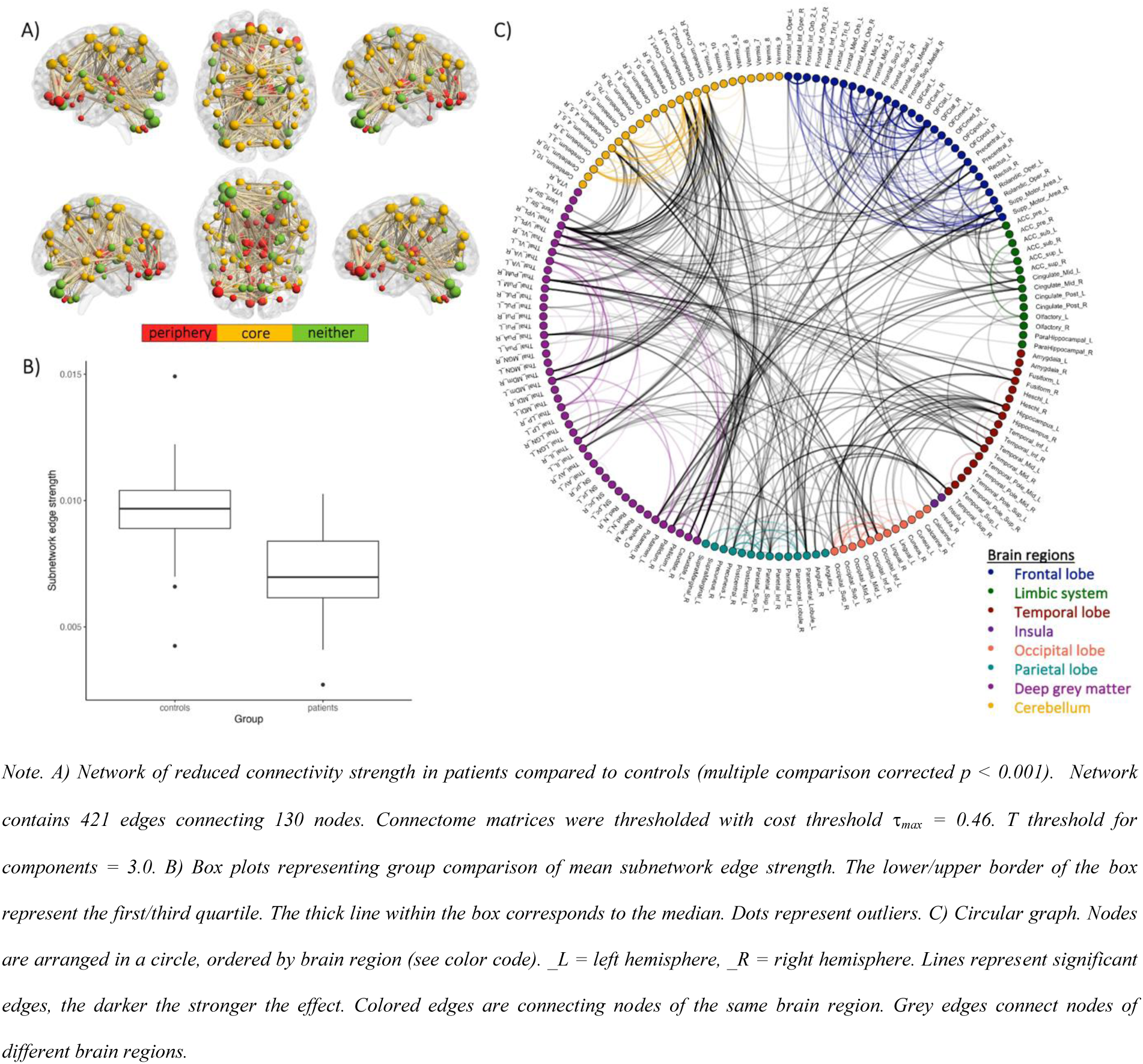
NBS-derived subnetwork with lower network connectivity strength in patients compared to controls at a T threshold of 3.0

### 3.3. Associations between network connectivity measures, clinical risk factors and family-environmental risk factors

Within the patient group, the CCR score was not significantly associated with global efficiency ((*β*(*CI-95*)=-0.26(−0.54–0.02),*p*=0.071). However, CCR score was significantly associated with local efficiency (*β*(*CI-95*)=-0.40(−0.67–-0.14),*p*=0.004) and subnetwork edge strength (*β*(*CI-95*)=-0.57(−0.79–-0.35),*p*<0.001).

Post hoc, a specific local subnetwork was identified with NBS, which was directly associated with the CCR score. This subnetwork consisted of 58 edges connecting 39 nodes (*p*<0.001, Figure 4, Supplementary table 3 and 6). This subnetwork included nodes of the frontal, parietal, and temporal, lobes, limbic system, insula, deep grey matter, cerebellum and was characterized by dense interconnections of the frontal lobe and strong fronto-parietal-thalamic connections. The majority of affected nodes were classified as core regions.

**Figure 4:**
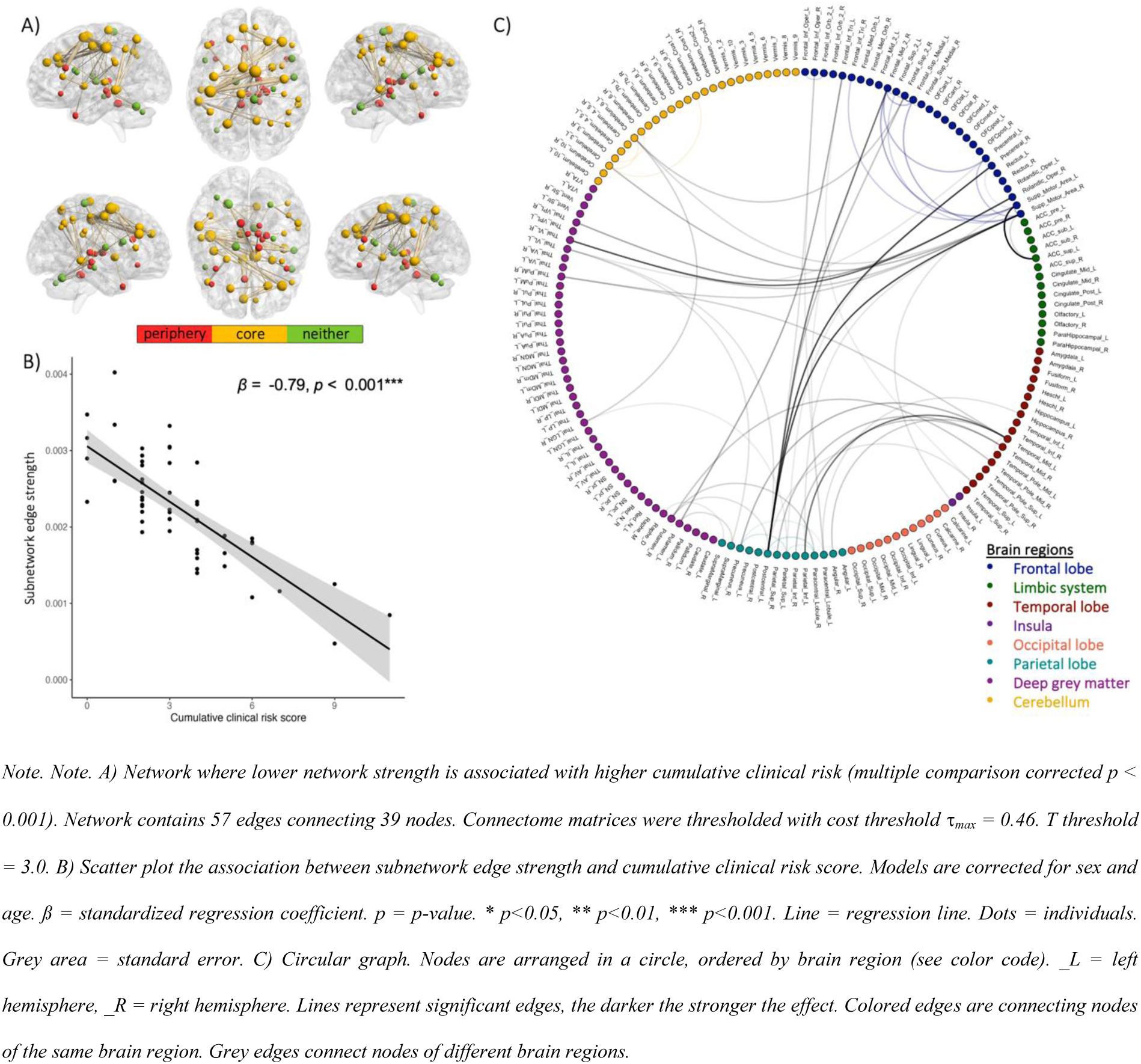
NBS-derived subnetwork where lower network connectivity strength is associated with higher cumulative clinical risk score

Post hoc associations with single risk factors were investigated. After FDR correction, longer length of ICU stay was significantly associated with lower local efficiency (*β*(*CI-95*)=-0.42(−0.69–-0.15),FDR-corrected *p*=0.034) and lower subnetwork edge strength (*β*(*CI-95*)=-0.53(−0.76–-0.30),*p*<0.001). Further, lower subnetwork edge strength was associated with higher number of heart medication (*β*(*CI-95*)=-0.40(−0.66–-0.16),*p*=0.005), higher number of CPB surgeries (*β*(*CI-95*)=-0.52(−0.74–-0.29),*p* < 0.001) and univentricular CHD (*β*(*CI-95*)=-0.53(−0.76–-0.30),*p*<0.001). See Figure 5 and Supplementary Table 7. We did not find an association between any structural network measures and family-environmental factors (all *p*>0.1, Supplementary Table 8).

**Figure 5:**
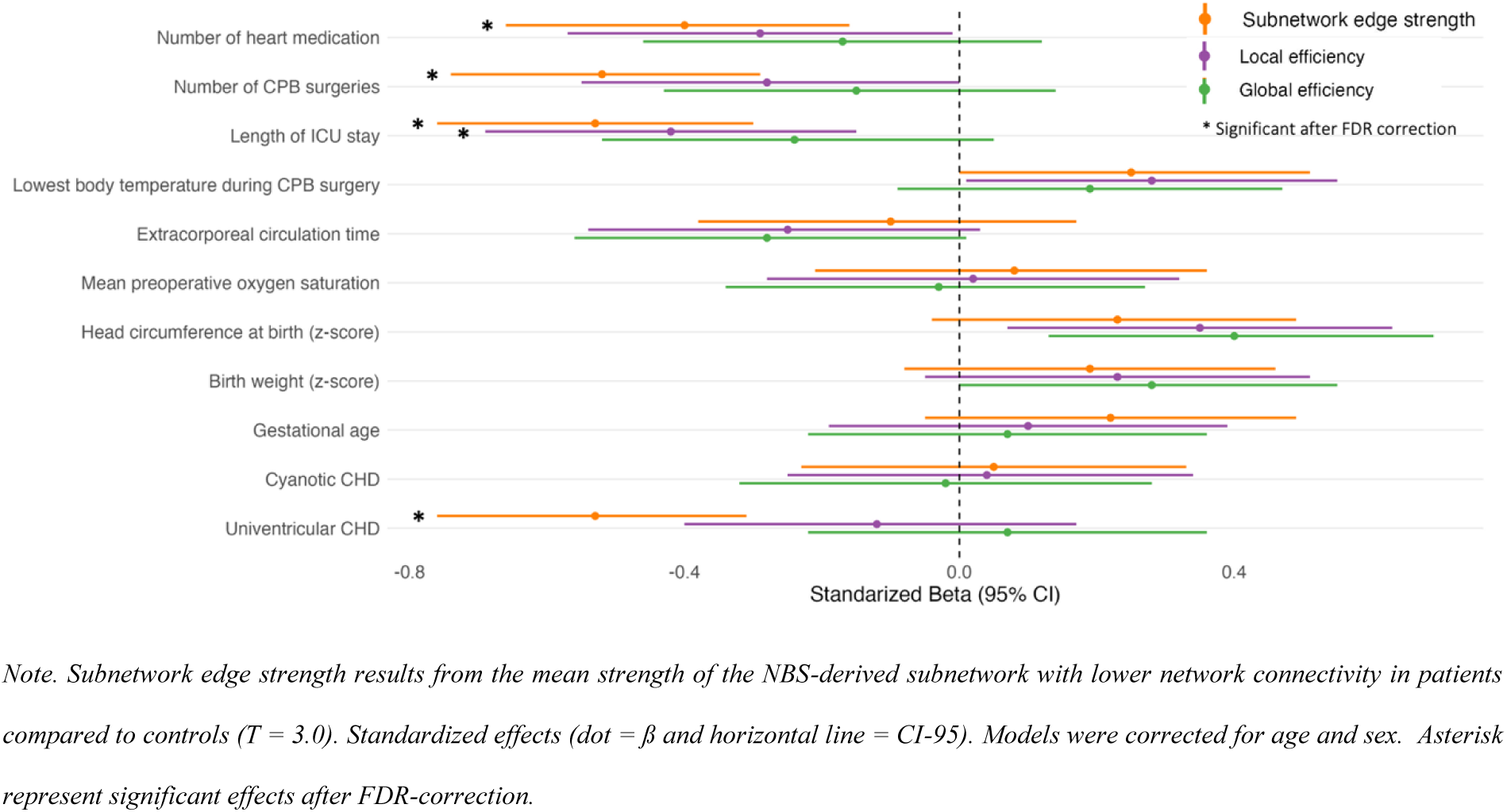
Forest plot representing the effect sizes of the associations between structural network features and single clinical risk factors

### 3.4. Associations between network connectivity measures and executive functions

The executive function summary score was significantly associated with global efficiency (*β*(*CI-95*)=0.34(0.07–0.61),*p*=0.015) and local efficiency (*β*(*CI-95*)=0.34(0.08–0.61),*p*=0.013) in patients. There was no significant association between the executive function summary score and subnetwork edge strength (*β*(*CI-95*)=0.24(−0.03–0.5),*p*=0.076). Post hoc, we tested if a specific local subnetwork was associated with executive function performance. However, by means of NBS no specific network was identified (*p*=0.380).

Lastly, we tested if the association between the CCR score and the executive function summary score was mediated by local efficiency. However, there was no significant mediation effect evident (*β=*-0.048,standard error=0.029,*z*-value=-1.627,*p*=0.104).

## 4. Discussion

In this study of adolescents with CHD, we showed that the cumulative exposure to clinical risk factors is associated with brain-wide alterations in structural connectivity. Specifically, we found lower network segregation and lower edge strength of a subnetwork including frontal interconnections and fronto-parietal-thalamic connections. Further, we showed that poorer executive functioning was associated with lower network integration and lower segregation, and on a trend-level with lower edge strength.

When comparing adolescents with CHD to healthy controls, we found that while network integration was preserved, adolescents with CHD showed lower network segregation and lower edge strength in a dense, widespread network connecting structures of all major brain regions (frontal, parietal, temporal, and occipital lobes, limbic system, insula, deep grey matter, cerebellum). Interconnections of the frontal lobe, interhemispheric connections and cortico-subcortical connections seem to be particularly affected by lower edge strength.

### 4.1. Clinical risk factors for altered network connectivity

This is the first study that investigated how structural brain networks are affected by the exposure to a cumulative burden of clinical risk factors. We found that a higher load of cumulative clinical risk (CCR) was associated with lower network segregation and lower edge strength in adolescence. A post hoc analysis showed that CCR was strongly associated with lower edge strength of a subnetwork involving frontal interconnections and fronto-parietal-thalamic connections.

While neurogenesis is largely complete by 20 weeks of gestation, the development of structural networks continues by means of axonal and dendritic growth, proliferation, followed by cell loss, synaptic pruning and axon myelination throughout childhood and adolescence.^38,39^ During this phase, patients with complex CHD undergo cardiac surgery and stay on the ICU, some of them have continued altered heart function and face severe cardiac and neurologic events (e.g., cardiac arrest, seizure, or stroke). It is likely that these continuous hits may further disrupt brain development and thus increase the extent of alterations in network connectivity over time.

Thalamocortical connections are established prenatally and develop strongly during the second and third trimesters.^34,35^ As this phase is marked by increased metabolic demands, altered cardiovascular physiology and decreased substrate delivery may prevent normal brain development of thalamocortical connections – particularly in those patients with more severe CHD.^36^ Across childhood and adolescence, structural networks are still maturing, especially the frontal cortex.^37^ During this time, patients with CHD may undergo surgeries, need to stay on the ICU and may even face severe neurologic or cardiac events (e.g., stroke, seizures, being on ECMO). The cumulation of these clinical risk factors over time may further impact the development of brain networks and alterations may become more evident. As mentioned above, alterations of structural networks seem to be stronger in adolescents with CHD compared to neonates with CHD and we have shown that the cumulation of clinical risk over time is linked to more network alterations. These findings indicate that structural brain networks remain vulnerable recuring clinical risk beyond the fetal and neonatal period.

We could show that creating a cumulative risk score using a set of predefined and previously identified risk factors is a valid and straight forward approach to predict both, alterations in structural brain networks and executive function impairments. Thus, such a risk score may serve as a biomarker and we suggest that this approach could be applied in other studies to predict neurodevelopmental outcome and brain development. Eventually a cumulative risk score may be used in clinical practice for outcome prediction, to evaluate the need for early interventions and to inform caregivers. However, it must be considered that while the effect sizes are moderate to strong, there remains a considerable amount of unexplained variance and thus uncertainty in prediction.

To further establish which individual risk factors may play a particular role for the development of structural brain networks, we conducted univariate linear regression models to investigate each risk factor separately. This analysis of single risk factors showed that prolonged stay on the ICU was the strongest predictor for lower network segregation and lower edge strength in adolescence. In addition, a univentricular CHD, more than one CPB surgery and a cardiac insufficiency determined by the need for cardiac medication at the time of assessment were also associated with lower edge strength.

The timing of risk factors may impact the structural network connectivity at different levels. Univentricular physiology may already impact intrauterine brain development including brain growth and the formation of fundamental networks such as thalamocortical connections. We could show that particularly network edge strength was associated with having a univentricular CHD while network integration and segregation were not associated with univentricular CHD.

After birth, the infant undergoes open-heart surgery and stays on the ICU which is known to be a stressful environment. We could show that long ICU stays were associated with lower network segregation and lower network edge strength. Cardiac intensive care strategies include modifiable variables and thus constitute a target to improve brain health in these children. Strategies to promote developmental care in the ICU may prevent or at least reduce the extent of brain alterations. These strategies can include promoting parent-child bonding with kangaroo care or music therapy, reducing noise and implementing cycled lighting to maintain the circadian rhythm.^40^

### 4.2. Altered global and local network connectivity in patients with CHD

Using graph theory to investigate global network connectivity, we demonstrated that adolescents with CHD had lower total network density and lower network segregation compared to healthy controls. In contrast to our findings, a previous study in adolescents with TGA showed significantly increased network segregation measured by modularity.^29^ This study used FA-weighted connectome data while we used sum of weighted streamlines generated by constrained spherical deconvolution (CSD). CSD enables the modelling of crossing fibers within voxels and thus, overcomes a major limitation of FA analyses.^41^ Further, the connectome data of this study was not cost corrected while our data was thresholded at different cost levels in order to eliminate false-positive connections and to prevent that the comparison of network connectivity is biased by the network density. The different results of the two studies could thus be attributed to methodological differences.^38^ However, future studies are needed to further establish the extent and direction of global topological differences between adolescents with CHD and healthy controls.

A body of research has demonstrated that structural brain alterations are already present during the prenatal and neonatal period. It has been suggested that reduced cerebral oxygen delivery, altered cerebral blood perfusion as well as placenta abnormalities in fetuses with CHD may disrupt the development of the vascular-rich subventricular zone, relevant for neurogenesis and oligodendrocyte myelination^42^, and of the fetal subplate, relevant for cortical maturation.^43^ Early disruption of these core regions may alter further development of structural networks throughout the fetal and neonatal period. Indeed, dMRI studies in neonates with CHD have demonstrated altered global network connectivity during the pre-and postoperative period. However, the specific findings were heterogenous and somehow contradicting with respect to the significance and direction of effects for network density, segregation and integration.^19–21^ To better understand different findings across dMRI studies, replicating processing pipelines and using meta-analytic approached should be used to summarize effect sizes and potential biases.

Using network-based statistics to investigate local network connectivity, we provided first evidence in adolescents with CHD for lower edge strength in a dense, widespread network including interconnections of the frontal lobe, interhemispheric connections and cortico-subcortical connections. Further, we found that connections of densely connected core regions as well as connections from and to the thalamic nuclei were affected by lower edge strength. Edges connecting periphery regions were less affected by lower edge strength. Previous studies in neonates with CHD have also demonstrated lower edge strength of subnetworks both pre and postoperatively.^20,21^ A qualitative comparison of the networks identified by these studies showed that effects were stronger and more wide spread in adolescents with CHD compared to neonates with CHD. This may indicate that network strength may be even more affected throughout development. Longitudinal studies are needed to confirm this hypothesis.

### 4.3. Association with executive function performance

We found that lower executive functioning in adolescents with CHD was associated with lower network integration and segregation. Our findings are in line with a previous study which investigated FA-weighted connectomes of 882 healthy individuals between 8 and 22 years of age. They found that, with increasing age, networks become more segregated, which was linked to better executive functioning.

They also found that networks become globally more integrated, indicating that the connectivity within modules is efficiently organized and hub edges build strong links between otherwise segregated modules.^44^

However, our findings are in contrast to a previous study in adolescents with TGA which found that higher network segregation, assessed by modularity, was associated with poorer executive functioning. As described previously, our results widely deviate from the findings of this study likely due to methodological differences. Additional studies are needed replicating previously conducted processing pipelines and statistical analyses to consolidate the evidence from current research.

While studies investigating associations between structural network connectivity and executive functioning remain sparse, additional studies have shown that poorer executive functioning is associated with locally altered microstructural integrity in adolescents and young adults with CHD.^45–47^ The use of different methodological approaches across studies prevents a direct comparison of results, but these studies repeatedly showed that patients with CHD demonstrate altered structural networks and microstructural integrity and that these alterations are linked to poorer executive functioning. Adolescence is a sensitive developmental period which is characterized by the reorganization of structural brain networks and maturation of white matter tracts. Brain plasticity, which plays a crucial role during this age period, has the potential to be promoted by specific therapeutic and interventional approaches. Initial studies have shown that computerized working memory programs in patients with CHD can improve executive functioning, though the improvement is limited in its extent.^48,49^ One study using a multimodal approach and combined weekly coaching sessions with a psychologist and a computerized cognitive training is currently being conducted.^50^ We are optimistic that future research using advanced imaging methodologies, longitudinal study designs, and testing executive function interventions will advance our understanding of neurological underpinnings of executive function impairments in patients with CHD.

### 4.4. Persisting brain alterations – a lifespan approach is needed

As elaborated above, we and others have shown that alterations in structural network connectivity in patients with CHD seem to persist from the neonatal period into adolescence. Also, we have shown previously in a pooled case-control study using diffusion tensor imaging that white matter microstructural alterations in patients with CHD persist throughout childhood into young adulthood.^27^ Importantly, the described brain alterations in patients with CHD are linked to executive function impairments which we frequently observe in this population. Due to these persisting brain alterations, the brain reserve of patients with CHD may be reduced. People who have sufficient brain reserve tolerate age-related changes better without showing clinically relevant symptoms of cognitive decline^51^. This is of particular importance when looking at a life span perspective. In healthy adults, natural age-related changes occur in the white matter by means of volume loss, microstructural changes in specific white matter tracts and changes in structural network connectivity most prominently in the frontal and temporal lobe. These changes are thought to build the structural substrate underlying cognitive decline in later adulthood.^52,53^ In the case of patients with CHD, persisting white matter alterations may reduce their brain reserve and additional age-related changes in the white matter may have amplified consequences. As brain reserve capacity is known to be protective for later dementia^51^, limited brain reserve in CHD may accelerate age-related cognitive decline. Indeed, studies have shown that adults with CHD have a 2-fold higher risk for early dementia.^54,55^ In addition, microvascular changes in adults with CHD may lead to accelerated vascular aging which can in turn increase the risk for brain alterations. Thus, a life span approach incorporating neuropsychologists and neurologists is highly needed when treating patients with CHD. Future studies are needed in elderly adults with CHD combining neuroimaging and cognitive assessments to better understand the neurocognitive and neurologic sequelae of these patients across the life span.

### 4.5. Limitations

Some limitations need to be considered when interpreting our results. We conducted a single-shell dMRI sequence with 35 diffusion-weighted gradient directions. Thus, the angular resolution of our dMRI data is limited. However, we applied constrained spherical deconvolution (SS3T-CSD) to estimate the fiber orientation distribution of each voxel.^41,56^ This approach enables the modelling of crossing fibers within voxels and thus, overcomes a major limitation of diffusion tensor imaging and improves the reliability of connectomic analysis.

We showed that different measures of altered network connectivity were associated with executive performance. However, it has to be mentioned that the data was collected cross-sectionally and no conclusion can be drawn on causality. Longitudinal studies are needed with repeated MRI examinations and neurodevelopmental follow ups to model developmental trajectories and infer causality.

The same limitation applies to environmental factors (i.e., parental mental health and family functioning), which were assessed cross-sectionally. While the current state of parental mental health and family functioning may not influence the adolescents’ brain networks directly, disrupted family well-being during early childhood may alter the trajectory of brain development. For instance, Wu and colleagues has shown that maternal depression during pregnancy is associated with reduced brain growth in fetuses with CHD.^57^ This is of particular importance as parents of children with CHD are at increased risk for mental health issues including posttraumatic stress disorder, depression and anxiety.^58^ Further longitudinal studies in this direction are highly needed.

Finally, the control group of this study population was a high-functioning group with a relatively high mean IQ of 111 and a high socioeconomic background. Also, the patients who participated in this study were from higher socioeconomic background, indicated by higher parental education, compared to those who refused to participate. Thus, group the generalizability of our results is limited.

### 4.6. Conclusion

In this study we used a novel dMRI approach, constrained spherical deconvolution, to model crossing fibers and estimate structural networks, in adolescents with CHD and healthy controls. By means of graph theory and network-based statistics, we demonstrated evidence for altered network connectivity in adolescents with CHD – most prominently in those patients who have faced a cumulative exposure to multiple clinical risk factors over time. This was the first study to investigate the cumulative load of risk in patients with CHD associated with structural brain networks and executive function performance. Quantifying the cumulative load of risk in patients with CHD may help to better predict trajectories of brain development. This would allow us to identify and support the most vulnerable patients as early as possible. Further, a better understanding of the role of clinical risk factors for brain development may inform clinical and surgical decision making.

## Data Availability

De-identified data and statistical scripts supporting our findings can be shared specifically with other groups upon reasonable request. Patient data cannot be made openly available due to patient privacy reasons.

## Abbreviations

BSI-18: Brief Symptom Inventory
CPB: cardiopulmonary bypass
CHD: congenital heart disease
CCR: cumulative clinical risk
dMRI: diffusion weighted MRI
ECC: extracorporeal circulation
ECMO: extracorporeal membrane oxygenation
FRI: family relationship index
ICU: intensive care unit
NBS: network-based statistics
SDS: standard deviation score
SPGR: spoiled gradient echo
SS3T-CSD: single-shell three-tissue constrained spherical deconvolution
TGA: transposition of the great arteries
TR/TE: repetition time/echo time

## Authors contribution

**ME** has acquired funding, participated in designing the study and data assessment, she has conducted the data processing, statistical analyses, interpretation of results and drafting of the manuscript. **AS** has conducted data processing and revised the manuscript. **OK** has participated in the interpretation of the data and revised the manuscript. **RT** has acquired funding, participated in designing the study and revised the manuscript. **BL** has acquired funding, participated in designing the study, provided supervision and revised the manuscript. **AJ** has acquired funding, participated in designing the study, provided supervision and revised the manuscript.

## Funding

This project was supported by the Swiss National Science Foundation (SNF 32003B_172914) and the University Research Priority Program (URPP) ‘Adaptive Brain Circuits in Development and Learning (AdaBD)’ of the University of Zurich. The sponsors had no influence on the study design, the collection, analysis, and interpretation of data, the writing of the manuscript, or the decision to submit the paper for publication.

We thank all participants and their families for participating in the study. We thank Thorsten Weirauch and the MRI team for their valuable support in performing the MRI scans. We thank Nadja Naef, Alenka Schmid, and Flavia Wehrle for their help in planning the study and acquiring the data. We thank Francois Mojon for his help in rating the registration quality. We thank Raimund Kottke for reviewing the MR images for incidental findings. We thank Valentin Rousson for his statistical advice.

